# Chloroquine diphosphate in two different dosages as adjunctive therapy of hospitalized patients with severe respiratory syndrome in the context of coronavirus (SARS-CoV-2) infection: Preliminary safety results of a randomized, double-blinded, phase IIb clinical trial (*CloroCovid-19 Study*)

**DOI:** 10.1101/2020.04.07.20056424

**Authors:** Mayla Gabriela Silva Borba, Fernando Fonseca Almeida Val, Vanderson Souza Sampaio, Marcia Almeida Araújo Alexandre, Gisely Cardoso Melo, Marcelo Brito, Maria Paula Gomes Mourão, José Diego Brito-Sousa, Djane Baía-da-Silva, Marcus Vinitius Farias Guerra, Ludhmila Abrahão Hajjar, Rosemary Costa Pinto, Antonio Alcirley Silva Balieiro, Felipe Gomes Naveca, Mariana Simão Xavier, Alexandre Salomão, André Machado Siqueira, Alexandre Schwarzbolt, Júlio Henrique Rosa Croda, Maurício Lacerda Nogueira, Gustavo Adolfo Sierra Romero, Quique Bassat, Cor Jesus Fontes, Bernardino Cláudio Albuquerque, Cláudio Tadeu Daniel-Ribeiro, Wuelton Marcelo Monteiro, Marcus Vinícius Guimarães Lacerda, CloroCovid-19 Team

**Author notes:** Correspondence to: Marcus Lacerda, Fundação de Medicina Tropical Dr. Heitor Vieira Dourado, Manaus, Amazonas, Manaus, 69040-000, Brazil.

## Abstract

**Background:** There is no specific antiviral therapy recommended for the disease caused by SARS-CoV-2 (COVID-19). Recent publications have drawn attention to the possible benefit of chloroquine (CQ). Our study aimed to comprehensively evaluate the safety and efficacy of two different CQ dosages in patients with established severe COVID-19.

**Methods:** We performed a parallel, double-blinded, randomized, phase IIb clinical trial, aiming to assess safety and efficacy of two different CQ dosages as adjunctive therapy of hospitalized patients with SARS in Manaus, Brazilian Amazon. Eligible participants were allocated to receive orally or via nasogastric tube high dose CQ (600mg CQ twice daily for 10 days or total dose 12g); or low dose CQ (450mg for 5 days, twice daily only on the first day, or total dose 2.7g). In addition, all patients received ceftriaxone and azithromycin. This study was registered with <u>ClinicalTrials.gov</u>, number NCT04323527.

**Findings:** Out of a pre-defined 440 patients sample size, 81 patients were enrolled. The high dosage CQ arm presented more QTc>500ms (18.9%), and a trend toward higher lethality (39%) than the lower dosage. Fatality rate until day 13 was 27% (95%CI=17.9-38.2%), overlapping with the CI of historical data from similar patients not using CQ (95%CI=14.5-19.2%). In 27 patients with paired samples, respiratory secretion at day 4 was negative in only six patients (22%).

**Interpretation:** Preliminary findings suggest that the higher CQ dosage (10-day regimen) should not be recommended for COVID-19 treatment because of its potential safety hazards. Such results forced us to prematurely halt patient recruitment to this arm. Given the enormous global push for the use of CQ for COVID-19, results such as the ones found in this trial can provide robust evidence for updated COVID-19 patient management recommendations.

**Funding:** This study was funded by the Government of the Amazonas State, *Farmanguinhos* (Fiocruz), SUFRAMA, CAPES, FAPEAM, and federal funds granted by a coalition of Brazilian senators.

**Research in context:** *Evidence before this study:* Before the CloroCovid-19 trial began, to our knowledge, there were no published reports of robust clinical studies on the safety and/or efficacy of chloroquine (CQ) and/or hydroxychloroquine (HCQ) for the treatment of COVID-19 during the recent 2020 pandemic. We searched PubMed and also MedRxiv.org (pre-print server for health sciences, without peer review), without any language restrictions and including Chinese publications, for studies published between Dec 2019 and April 5, 2020, using the search terms ‘COVID-19, coronavirus, SARS-Cov-2’. We found three non-randomized studies with limited sample sizes in which (1) HCQ use led to a decrease in SARS-Cov-2 detected in respiratory secretions five days after treatment, together with azithromycin (France, 36 patients); (2) HCQ use shortened time to clinical recovery (China, 62 patients); and (3) CQ was superior to control treatment in inhibiting the exacerbation of pneumonia, improving lung imaging findings, and promoting virus-negative conversion and shortening the disease course (China, 100 patients). We found no published studies comparing different dosages of CQ/HCQ and their thorough safety assessment.

*Added value of this study:* In a larger patient population, we found that a higher dosage of CQ for 10 days presented toxicity red flags, particularly affecting QTc prolongation. The limited sample size recruited so far does not allow to show any benefit regarding treatment efficacy, however the higher fatality associated with the higher dosage by day 13 of follow-up resulted in a premature halting of this arm. This is the first double-blinded, randomized clinical trial addressing different dosages of CQ for the treatment of severe patients with COVID-19 in the absence of a control group using placebo. Due to the impossibility of not using the drug recommended at the national level, we used historical data from the literature to infer comparisons for lethality endpoints. Follow-up until day 28 is ongoing with a larger sample size, in which long-term lethality will be better estimated.

*Implications of all the available evidence:* The preliminary findings from CloroCovid-19 trial suggest that the higher dosage of CQ (12 g total dose over 10 days) in COVID-19 should not be recommended because of safety concerns regarding QTc prolongation and increased lethality, in the Brazilian population, and more often in older patients in use of drugs such as azithromycin and oseltamivir, which also prolong QTc interval. Among patients randomized to the lower dosage group (5 days of treatment, total dose 2.7 g), given the limited number of patients so far enrolled, it is still not possible to estimate a clear benefit of CQ in patients with severe ARDS. Preliminary data on viral clearance in respiratory secretions in our confirmed cases are also indicative of little effect of the drug at high dosage. More studies initiating CQ prior to the onset of the severe phase of the disease are urgently needed.

## INTRODUCTION

Coronaviruses, first discovered in the 1960s, are a family of RNA viruses that typically cause respiratory and intestinal infections in birds and mammals. In humans, coronaviruses often cause mild upper respiratory tract infections, and together with rhinoviruses are the two main underlying aetiologies for the normal cold, with severe disease secondary to these viruses usually restricted to immunocompromised individuals^1^. In 2002, however, and as a result of a coronavirus-associated outbreak of severe acute respiratory syndrome (SARS), a pathogenic role was established^2^. This first SARS-coronavirus (SARS-CoV) outbreak appeared in south- eastern China and Hong Kong and quickly spread to various parts of the world, highlighting its pandemic potential and leading to significant economic losses^3,4^. A decade later, in 2012, a second highly pathogenic coronavirus, the Middle Eastern respiratory syndrome coronavirus (MERS-CoV), emerged in countries in the Middle East^5^. The virus was first isolated in June 2012^5^. By the end of 2016, more than 1850 cases of laboratory-confirmed MERS-CoV had been documented, with a case fatality rate of 35%^6^.

The first cases of the new coronavirus 2019 disease (COVID-19) were reported in December 2019, when a group of patients was admitted to hospitals in Wuhan, the capital of the Hubei province in Central China, with an initial diagnosis of pneumonia of unknown etiology^7^. Initially the outbreak with the new SARS-CoV-2 coronavirus (coronavirus disease 2019; formerly 2019-nCoV), was confined to the Hubei province, but it rapidly spread to many other countries^8,9^, compelling WHO to officially declare a global pandemic on March 11, 2020. The origin of the virus has yet to be fully elucidated, but genomic analysis suggests that it is closely related to viruses previously identified in bats^10^.

SARS-CoV-2 infection appears to cause a wide range of symptoms, encompassing asymptomatic infection, mild infections of the upper respiratory tract, severe viral pneumonia, respiratory failure, multiple organ failure and more deaths than previously expected^11^. Some studies have shown detailed clinical features of patients with SARS-CoV-2-associated viral pneumonia (SARS-CoV-2 pneumonia)^12^. Of laboratory confirmed patients in China, 5% had critical illnesses and almost 50% of the critical patients died, with an overall rate of fatal cases (2.3%) estimated to be about ten-fold higher than that observed for seasonal influenza^13^. Most deaths involved older adults, many of whom had underlying chronic diseases^14,15^.

Currently, there is no specific antiviral therapy recommended for coronavirus infections. Few treatment studies have been carried out because most strains of human coronavirus cause self-limiting disease, and routine supportive care is usually effective. For past severe strains of coronavirus, outbreaks were scattered, thus not allowing timely clinical trials. Since the 2002 SARS outbreak, new therapeutic agents targeting viral entry pathways, proteins, proteases, polymerases and methyltransferases have been tested in randomized clinical trials, with little success. Recent publications have drawn attention to the possible benefit of chloroquine sulphate and phosphate salts (chloroquine diphosphate-CQ) and hydroxychloroquine (HCQ) for the treatment of SARS-CoV-2 infected patients^16–21^. Both drugs historically have been used for the treatment of acute malaria, as well as in some chronic rheumatic conditions. HCQ, a derivative of CQ first synthesized in 1946, proved to be less (∼40%) toxic when used for longer periods of time than the three-day course recommended for malaria. HCQ is therefore one of the drugs recommended for the treatment of systemic lupus erythematosus and rheumatoid arthritis^22^. Although both drugs have a bitter taste, they are generally very well tolerated, and after millions of doses used, their accumulated safety database is massive. In prolonged use (months or even years), which is not the targeted scenario in COVID-19, CQ may deposit in many tissues, especially the eye, causing retinal toxicity^23,24^. Myopathy has also been associated with the use of CQ^25^. The major complication, even in short regimens, is the potential for QTc prolongation, favoring fatal arrhythmias such as ventricular tachycardia and *torsades de pointes*^26^.

The *in vitro* antiviral activity of QC was first identified in the late 1960s^27,28^. Two studies have shown anti-SARS-CoV activity^17,19^. Several studies suggest that CQ and HCQ have potential broad-spectrum antiviral activity, result in an increase in the endosomal pH required for virus/cell fusion, interfere with the glycosylation of SARS-CoV cell receptors and have anti-viral, anti-inflammatory and immunomodulating effects that together may provide effective treatment of patients with COVID-19 pneumonia^19,29,30^.

In 100 COVID-19 affected patients, the effect of CQ was superior to the control treatment in inhibiting the exacerbation of pneumonia, improving pulmonary imaging findings and promoting a negative conversion of the virus and reducing the disease course^20^. Gautret et al.^21^ evaluated 20 COVID-19 patients treated with 200 mg HCQ three times per day for ten days. Six patients also received azithromycin. The proportion of patients who tested negative in nasopharyngeal samples differed significantly between treated patients and controls on days 3-4-5 and 6 after inclusion. On day 6 after inclusion, 100% of patients treated with a combination of HCQ and azithromycin were considered ‘virologically cured’ compared with only 57.1% in patients treated with HCQ alone and 12.5% in the control group. These results, albeit highly preliminary and probably not sufficiently powered to be conclusive, supported an effort to evaluate more thoroughly the effect of CQ in the evolution and prognosis of COVID-19.

*Health Commission of Guangdong Province*^18^ recommended the use of CQ tablets at a dose of 500 mg twice daily for 10 days (total dose 10g), for the treatment of patients aged 18-65 years with mild, moderate or severe pneumonia secondary to COVID-19, as long as there were no specific contraindications. However, to guarantee an adequate patient follow-up, a strict monitoring and evaluation plan for the safety and efficacy is recommended. As opposed to the 10-day treatment recommended and evaluated in different studies, CDC^31^ initially recommended for adults a loading dose consisting of 600 mg of CQ base (6 tablets of 100 mg), followed by 300 mg after 12 h on day 1, then 300 mg bid, given orally on days 2 to 5. This shorter treatment regimen (5 versus 10 days) would potentially reduce the side effects and assumes a drug half-life of about 30 hours.

The fact that in many countries the ‘compassionate use’ of CQ or HCQ has already been formally indicated for severe patients, made it unethical to test proper efficacy due to the lack of a placebo arm as a comparator. Our study aimed to comprehensively evaluate primarily the safety, and secondarily the efficacy of CQ in two different dosages, as compared to historical data reported in the literature for similar severe patients not receiving CQ for the treatment of severe respiratory syndrome caused by COVID-19. Here, we report the data of the first 81 randomized patients.

## Methods

### Ethical aspects

This study was conducted in accordance with the principles of the Declaration of Helsinki and the Good Clinical Practice guidelines of the International Conference on Harmonization.

The protocol was timely approved by the *Brazilian Committee of Ethics in Human Research* (CONEP approval 3.929.646/2020). All patients and/or legal representatives in case of unconsciousness, were informed about objectives and risks of participation. They were given time to carefully read and then sign an informed consent form (ICF). After recovery, the patient also signed the ICF. Random online clinical monitoring and quality control was performed. A virtual independent *Data Safety and Monitoring Board* (DSMB), with epidemiologists, clinicians and experts in infectious diseases, was timely implemented to review the protocol and with daily meetings to follow-up the activities of the study. The trial was reported according to *Consolidated Standards of Reporting Trials* (Consort) statement.^32^

### Study design and site

CloroCovid-19 was a parallel, double-blind, randomized, phase IIb clinical trial, which started on March 23^rd^, 2020, aiming to assess safety and efficacy of CQ in the treatment of hospitalized patients with severe respiratory syndrome secondary to SARS-CoV-2 infection (ClinicalTrials.gov, number NCT04323527).

This trial is being conducted at *Hospital e Pronto-Socorro Delphina Rinaldi Abdel Aziz*, in Manaus, Western Brazilian Amazon (currently the biggest public reference unit dedicated exclusively to the treatment of severe COVID-19 cases in Brazil, with capacity to hospitalize 350 patients in Intensive Care Units - ICU). The hospital has all source documents registered on-line in an electronic medical recording system (*Medview*). Clinical analyses laboratory and routine CT scanning are also available locally.

Manaus is the capital of the Amazonas State, the biggest Brazilian State, and has ∼2.5 million inhabitants scattered in the ninth largest country subdivision of the world (>1.5M/km^2^). It is a major industrial, academic and tourist centre in the Amazon region, with several transportation hubs and thousands of annual foreign visitors. It is mostly served by the socialized and free *Unified Health System* (SUS) in an organized health assistance network, but also counts with many private hospitals. The city also counts with various universities, graduate programs and traditional clinical research groups dedicated to the study of infectious diseases. At the beginning of the study, autochthonous SARS-CoV-2 transmission had already been recorded at the study site.

### Participants

Hospitalized patients aged 18 years or older at the time of inclusion, with respiratory rate higher than 24 rpm AND/OR heart rate higher than 125 bpm (in the absence of fever) AND/OR peripheral oxygen saturation lower than 90% in ambient air AND/OR shock (defined as mean arterial pressure lower than 65 mmHg, with the need for vasopressors medicines or oliguria or a lower level of consciousness) were included. Children under 18 years of age were not included due to the known lower morbidity/mortality from COVID-19^33^. Patients were enrolled before laboratorial confirmation of COVID-19, considering that such procedure could delay randomization. For the analyses at this point, all patients were included regardless of the confirmed etiology which for safety issues (the focus of this manuscript) should not be an issue. For now, the flowchart of the study presents clinical-epidemiological suspected cases and cases already confirmed by RT-PCR.

### Sample size calculation

The sample for the primary outcome (reduction in lethality) was calculated assuming a 20% lethality incidence in critically ill patients not using CQ (historical control)^15,34,35^, and that both arms of CQ would be equally able to reduce lethality by at least 50%. Thus, considering a test of differences in proportions between 2 groups of the same size, 80% power and 5% alpha, 394 participants were needed (197 per group). Adding 10% of losses, the final sample of 440 participants was obtained. Sample calculation was performed in the R statistical package (v3.6.1), with the functions implemented in the TrialSize and gsDesign packages.

### Procedures

The interventions tested in this study were based on different regimens using CQ 150mg tablets (*Farmanguinhos*, Fiocruz, Brazil). Eligible participants were allocated at a 1:1 ratio to receive orally (or via nasogastric tube in case of orotracheal intubation) either: a) high dosage CQ (600mg CQ (4×150mg tablets, twice daily for 10 days, **total dose 12g**); or b) low dosage CQ (450mg CQ (3×150mg tablets + 1 placebo) twice daily on day 0, 3×150mg tablets +1 placebo tablet followed by 4 placebo tablets from D1 to D4, and then 4 placebo tablets twice daily from D5-D9, **total dose 2**.**7g**). Placebo tablets also produced by *Farmanguinhos* were used in the latter in order to standardize treatment and blinding of research team and participants.

As per hospital protocol, all patients meeting the same criteria of the study (ARDS) used intravenous ceftriaxone (1g 2x for 7 days) plus azithromycin (500mg 1x for 5 days), systematically, starting on day 0. Oseltamivir (75mg 2x for 5 days) was also prescribed when influenza infection was suspected (in the Amazon, the ongoing flu season is from January-April).

Clinical parameters were measured daily by the routine clinical staff from day 0 to discharge or death, and then at days 14 and 28 for discharged patients, to assess efficacy and safety outcomes. Laboratorial parameters and ECG were performed whenever needed at clinical discretion. Data were recorded on *Medview* and then transferred into an electronic database (REDCap), in tablet computers, at bedside in the wards, further validated by external trial monitoring staff.

### Outcomes

Safety outcomes included adverse events (AE) that occurred during treatment, serious adverse events (SAE), and premature or temporary discontinuation of treatment. Adverse events were classified according to the *National Cancer Institute Common Terminology Criteria for Adverse Events*. The working hypothesis around which this trial was designed was the halving of mortality in both groups by day 28. Thus, the primary endpoint was mortality by D28. Secondary endpoints included mortality on day 13, participant’s clinical status, laboratorial exams, and ECG on days 13 and 28, daily clinical status during hospitalization, duration of mechanical ventilation (if applicable) and supplementary oxygen (if applicable), and the time (in days) from treatment initiation to death. Here we present only analyses until day 13. A subgroup of patients enrolled when already admitted to ICU was analysed separately. Virologic measures included viral RNA detection on days 0 and 4.

### Randomization and masking

An electronically generated randomization list was prepared by an independent statistician, with four blocks of 110 participants per block. This randomization list associated each patient’s study number with an opaque surface hiding the treatment group designation. The list was accessible only to non-blinded pharmacists in the study, in an attempt to minimize observation bias. Participants were randomized by the study pharmacist to their designated treatment regimen at the time of inclusion and were subsequently identified throughout the study only by their allocated study number, always assigned following chronological order. In case of SAEs, unmasking was available to DSMB members, and preliminary analyses were performed even before the scheduled interim analyses, in order to guide early halting of any of the CQ arms.

### Laboratory

Hematology and biochemistry analyses were performed in automatized machines. Samples (from two nasopharyngeal or one oropharyngeal swabs) were submitted to viral RNA extraction using QIAamp Viral RNA Mini Kit Viral RNA mini kit, according to the manufacturer’s recommendations. Subsequently, all specimens for SARS-CoV-2 were tested using the protocol developed by the *US Centers for Disease Control and Prevention* (CDC/USA), updated on March 15, 2020 (https://www.fda.gov/media/134922/download), targeting the virus nucleocapsid (N) gene and the human RNase P gene, as an internal control. For all assays, specimens were considered positive if both viral targets, N1 and N2, showed Ct lower than 40.00. No quantitative RT-PCR data were presented here. Swab specimens were collected on D0 and D4.

### Statistical analysis

An intention-to-treat analysis was conducted as part of the primary safety and efficacy analysis. Untaken or mistaken tablets, and dosage correction pending on renal and liver failure were not systematically registered daily, not allowing therefore per protocol analysis. Descriptive statistics were used for demographic, laboratory and clinical data. To assess the safety of the high and the low dosages of CQ the proportion (95% CI) of deaths in each group was compared with the historical proportion (95% CI) of deaths in patients who did not use CQ in other countries. For qualitative variables, Chi-square tests and Fisher’s exact test were performed. t-test or Mann-Whitney test were used for means/median comparisons. An accumulated proportion of detection was assessed by survival models, using Kaplan-Meier estimate curves. Log-rank and Peto-Peto (correction for low observation numbers in the end of the follow-up) tests were used for survival time to event analyses. Statistical analyses were performed in the R statistical package (v3.6.1), and p<0.05 was considered significant.

## Results

### Population characteristics

At the study site, 81 patients were randomized as per protocol (41 in the high dosage CQ arm and 40 in the low dosage CQ arm; Figure 1 flowchart). Most patients were confirmed COVID-19 by RT-PCR *a posteriori* (62/81, 76.5%). The non-confirmed patients presented compatible clinical and epidemiological COVID-19 presentation, and were analysed together.

**Figure 1.**
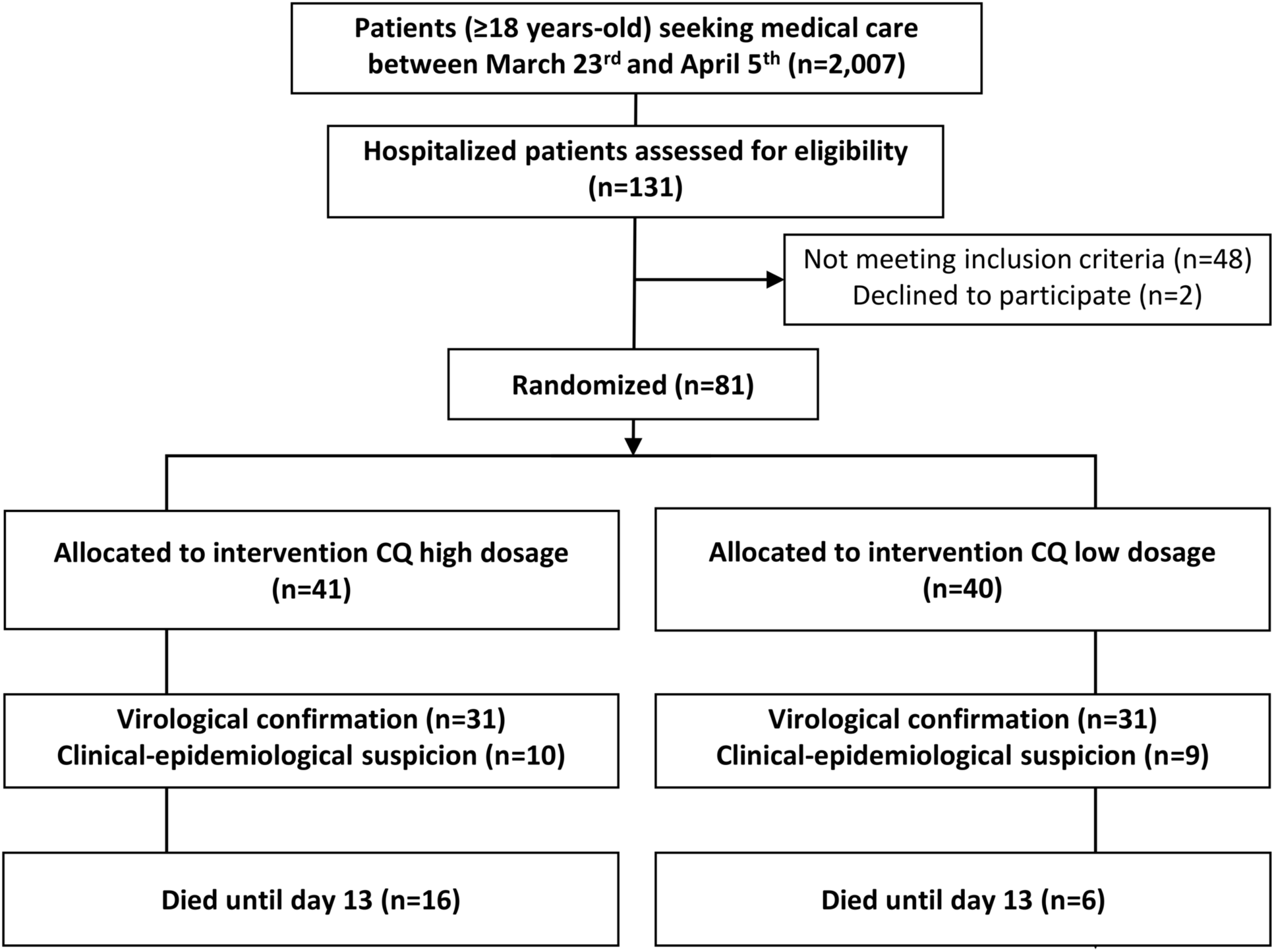
Study flowchart. Eligible participants were allocated at a 1:1 ratio to receive CQ to two arms at either high dose (600g CQ twice daily for 10 days) or low dose CQ (450mg CQ twice daily on the first day and 450mg once a day for the remaining 4 days, for a total of 5 days).

Older patients (aged over 75) were only enrolled in the high dosage CQ arm (n=5; Table 1). All the other characteristics were similar between age groups, allowing proper comparison. History of heart disease was more frequent among patients receiving the higher CQ dosage (p=0.05). Occurrence of myocarditis (defined as CKMB higher than 2x the upper normal limit), which may be a final complication of severe sepsis or a lesion triggered by the virus itself, was seen in 2/24 (8.3%) patients (1 patient/arm). No echocardiogram was performed timely.

**Table 1.** Demographic, clinical, laboratory, and radiographic findings of patients at baseline.

| **Variable** | **Total** | **CQ low dosage¶** | **CQ high dosage§** | **p-value** |
| --- | --- | --- | --- | --- |
| **Age, years, mean (SD)** | 51.1 (13.9) | 47.4 (13.3) | 54.7 (13.7) | **0.02** |
| **Age groups, years, %** |  |  |  |  |
| 18-50 | 41/81 (50.6) | 24/40 (60.0) | 17/41 (41.5) | **0.04** |
| >50-75 | 35/81 (43.2) | 16/40 (40.0) | 19/41 (46.3) |  |
| >75 | 5/81 (6.2) | 0/40 (0) | 5/41 (12.2) |  |
| **Female, %** | 20/81 (24.7) | 10/10 (25.0) | 10/41 (24.4) | 0.94 |
| **Race, %** |  |  |  |  |
| White | 17/81 (21.0) | 10/40 (25.0) | 7/41 (17.1) | 0.53 |
| Admixed | 58/81 (71.6) | 28/40 (70.0) | 30/41 (73.2) |  |
| Black | 6/81 (7.4) | 2/40 (5.0) | 4/41 (9.8) |  |
| **Health professional, %** | 5/81 (6.2) | 1/40 (2.5) | 4/41 (9.8) | 0.36 |
| **Pregnancy, %** | 2/20 (10.0) | 1/10 (10.0) | 1/10 (10.0) | 1.00 |
| **History of smoking, %** |  |  |  |  |
| Never smoked | 33/48 (68.8) | 18/24 (75.0) | 15/24 (62.5) | 0.21 |
| Current smoker | 4/48 (8.3) | 3/24 (12.5) | 1/24 (4.2) |  |
| Former smoker | 11/48 (22.9) | 3/24 (12.5) | 8/24 (33.3) |  |
| **Comorbidities, %** |  |  |  |  |
| Hypertension | 25/55 (45.5) | 10/27 (37) | 15/28 (53.6) | 0.28 |
| Diabetes | 14/55 (25.5) | 5/27 (18.5) | 9/28 (32.1) | 0.50 |
| Alcoholism | 14/51 (27.5) | 8/26 (30.8) | 6/25 (24) | 0.84 |
| Heart disease | 5/55 (9.1) | 0/27 (0) | 5/28 (17.9) | 0.06 |
| Asthma | 4/54 (7.4) | 1/26 (3.8) | 3/28 (10.7) | 0.61 |
| Chronic kidney disease | 4/54 (7.4) | 1/26 (3.8) | 3/28 (10.7) | 0.36 |
| Rheumatic diseases | 3/55 (5.5) | 3/27 (11.1) | 0/28 (0) | 0.17 |
| Liver diseases | 2/55 (3.6) | 2/27 (7.4) | 0/28 (0) | 0.24 |
| Tuberculosis | 2/55 (3.6) | 2/27 (7.4) | 0/28 (0) | 0.43 |
| HIV/Aids | 1/55 (1.8) | 0/27 (0) | 1/28 (3.6) | 0.48 |
| **Influenza vaccine in the last two years, %** | 16/81 (19.8) | 8/40 (20.0) | 8/41 (19.5) | 0.97 |
| **Medicines on admission, %** |  |  |  |  |
| Corticoid anti-inflammatories | 4/62 (6.5) | 3/32 (9.4) | 1/30 (3.3) | 0.61 |
| ACE inhibitors | 6/65 (9.2) | 2/34 (5.9) | 4/31 (12.9) | 0.29 |
| Bronchodilators | 2/78 (2.6) | 1/38 (2.6) | 1/40 (2.5) | 1.00 |
| **Oxygen therapy on admission, %** | 72/81 (88.9) | 36/40 (90) | 36/41 (87.8) | 1.00 |
| **Days from illness onset to hospital admission, median (IQR)** | 7 (4,9) | 6.5 (4,9) | 7 (5,10) | 0.82 |
| **Temperature distribution, %, °C** |  |  |  |  |
| <37.5 | 59/79 (74.7) | 30/39 (76.9) | 29/40 (72.5) | 0.46 |
| 37.5-38.0 | 10/79 (12.7) | 6/39 (15.4) | 4/40 (10) |  |
| 38.1-39.0 | 10/79 (12.7) | 3/39 (7.7) | 7/40 (17.5) |  |
| **Heart rate**, bpm, mean (SD) | 91 (17.5) | 91.6 (18.7) | 90.4 (16.4) | 0.76 |
| **Respiratory rate**, rpm, median (IQR) | 26.0 (21.0,30.0) | 25.0 (22.0,30.0) | 28.0 (20.0,31.0) | 0.53 |
| **Mean blood pressure**, mmHg, mean (SD) | 94.4 (17.1) | 96.2 (18.8) | 92.7 (15.4) | 0.38 |
| **Body mass index**, kg/m<sup>2</sup>, median (IQR) | 28.1 (26.0,31.6) | 28.9 (26.1,32.7) | 27.1 (25.7,31.2) | 0.30 |
| **Capillary refill time**, seconds, % | 13/55 (23.6) | 6/26 (23.07) | 7/27 (25.9) | 1.00 |
| **Oxygen saturation (SpO<sub>2</sub>)**, %, median (IQR) | 96 (94.0,98.0) | 96 (93.0,98.0) | 95 (94.0,98.2) | 0.91 |
| **White blood cell count**, ×10<sup>9</sup>/L, mean (SD) | 10.1 (4.6) | 10 (4.5) | 10.2 (4.8) | 0.88 |
| **Hemoglobin**, g/L, mean (SD) | 12.8 (2.3) | 13.2 (2.6) | 12.4 (1.9) | 0.20 |
| **Platelet count**, ×10<sup>9</sup>/L, median (IQR) | 211.0 (182.8,258.5) | 196.5 (172.5,256) | 215.0 (184.2,257.5) | 0.66 |
| **Alanine aminotransferase**, U/L, median (IQR) | 65.2 (49.7,103.8) | 51 (39.1,53.8) | 100 (92.3,115.1) | 0.05 |
| **Creatinine**, μmol/L, median (IQR) | 1.5 (1.1,2.7) | 1.3 (1.0,2.2) | 1.6 (1.1,3.0) | 0.25 |
| **Lactate dehydrogenase**, U/L, median (IQR) | 948 (810.0,1139.8) | 900 (553.0,1009.0) | 1010 (869.0,1337.5) | 0.39 |
| **Creatine kinase**, U/L, median (IQR) | 95.2 (61.9,250.4) | 82.8 (55.8,177.4) | 96.8 (70.8,279.0) | 0.39 |
| **Creatine kinase MB**, U/L, median (IQR) | 20 (15.8,25.9) | 18.6 (15.8,24.5) | 20.9 (15.8,27.3) | 0.83 |
| **International Normalized Ratio**, mean (SD) | 1.1 (0.1) | 1.1 (0.1) | 1.2 (0.1) | 0.40 |
| **C-reactive protein**, mg/L, median (IQR) | 84.8 (69.8,94.7) | 80.9 (61.9,95.1) | 86.1 (77.3,91.9) | 0.29 |
| **QTc**, ms, mean (SD) | 424.7 (27.4) | 421.9 (24.0) | 427.8 (31.0) | 0.49 |
| **Radiological findings**, % |  |  |  |  |
| Unilateral ground-glass opacity infiltration | 41/81 (50.6) | 20/40 (50.0) | 21/41 (51.2) | 0.91 |
| Bilateral ground-glass opacity infiltration | 8/81 (9.9) | 6/40 (15.0) | 2/41 (4.9) | 0.16 |
| Unilateral consolidation | 25/81 (30.9) | 15/40 (37.5) | 10/41 (24.4) | 0.20 |
| Bilateral consolidation | 15/81 (18.5) | 7/40 (17.5) | 8/41 (19.5) | 0.82 |
| Pleural effusion | 5/81 (6.2) | 3/40 (7.5) | 2/41 (4.9) | 0.66 |
| **qSOFA score <2**, % | 54/81 (66.7) | 30/40 (75.0) | 24/41 (58.5) | 0.12 |

§ High dose CQ (600g CQ twice daily for 10 days) for 10 days;
¶ Low dose CQ for 5 days (450mg CQ twice daily on the first day and 450mg once a day for the remaining 4 days).
For some variables, patients unconsciousness did not allow for complete personal history data collection.

### Safety outcomes

One patient developed severe rhabdomyolysis, and causality could be attributed to the virus or to CQ, which is already known to cause myolysis (Table 2). Two patients in the high dosage CQ arm evolved with ventricular tachycardia before death. This severe type of arrythmia is usually facilitated when QTc is prolonged.

**Table 2.** Safety outcomes in the intention-to-treat population until Day 13*.

| Variable | Total | CQ low dosage¶ | CQ high dosage§ | *p*-value |
| --- | --- | --- | --- | --- |
| Hemoglobin decreased¥, % | 11/42 (26.2) | 4/18 (22.2) | 7/24 (19.2) | 0.73 |
| Creatinine increased‡, % | 16/38 (42.1) | 7/15 (46.7) | 9/23 (39.1) | 0.74 |
| Creatine phosphokinase (CK) increased | 13/33 (39.4) | 6/19 (31.6) | 7/14 (50.0) | 0.47 |
| CKMB increased | 10/26 (38.4) | 3/13 (23.1) | 7/13 (53.8) | 0.23 |
| QTcF >500ms‡, % | 11/73 (15.1) | 4/36 (11.1) | 7/37 (18.9) | 0.51 |
| Ventricular tachycardia* (%) | 2/73 (2.7) | 1/36 (2.8) | 2/37 (5.4) | 0.51 |

\* Not all patients have completed Day 13 visit until this publication was finalized
¶ Low dose CQ for 5 days (450mg CQ twice daily on the first day and 450mg once a day for the remaining 4 days);
§ High dose CQ (600g CQ twice daily for 10 days).
¥ Shown are decreases in hemoglobin level of more than 3 g per deciliter or 30% or more from baseline.
¶ Shown are increases in serum levels of 30% or more from baseline.
‡ Severe adverse events related to the trial regimen were prolongation of the QT interval corrected for heart rate according to Fridericia's formula (QTcF).

No differences in hematological or renal toxicity was seen between the groups.

### Efficacy until day 13 outcomes

Of 27 patients with paired samples (in both arms), respiratory secretion at day 4 was negative in only six patients (22%).

The fatality rate in our sample was 27% (95%CI=17.9-38.2%), therefore still overlapping with the CI of the meta-analysis based on two major studies, which used similar patients without CQ (95%CI=14.5-19.2%). Survival per arm was presented in comparison to historical collation of available data from two other similar lethality studies with patients not receiving CQ (Figure 2). Both arms were very similar to these data showing no clear differences, despite more deaths in the higher dosage CQ arm (p=0.03). A subgroup was analysed with critically-ill patients enrolled, and compared to the large historical sample-size cohort of patients in Lombardy, Italy (Figure 3). No differences were seen between groups, but fatality seemed to be higher than in Italy, where patients were not using CQ. Only two out of 22 deaths were in older than 75 years-old. Nineteen out of 22 deaths had virological confirmation *antemortem*.

**Figure 2.**
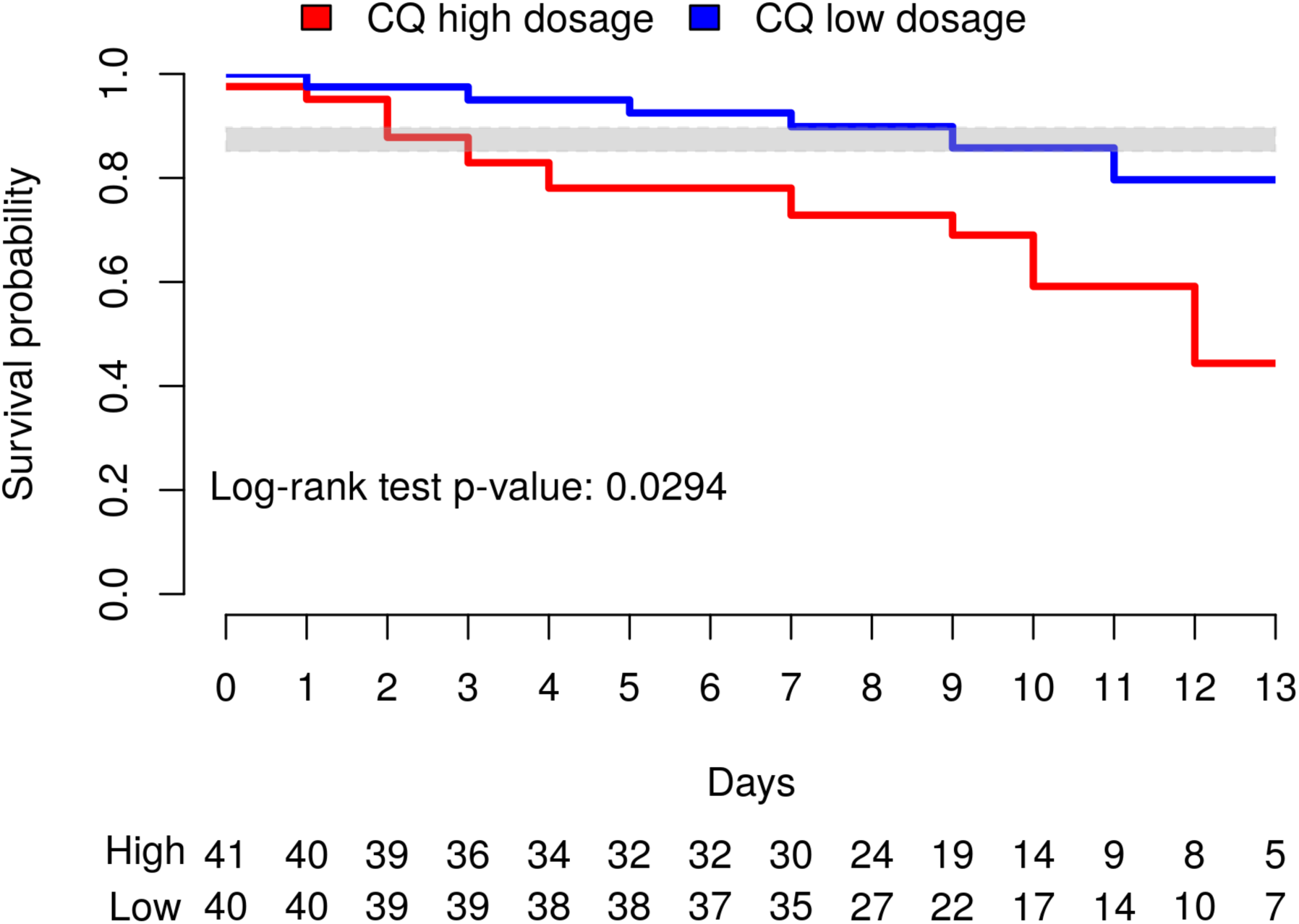
Time (in days) from randomization to death, in patients treated with each chloroquine dosage. The gray band represents the upper and lower limits of the confidence interval for lethality in hospitalized patients not receiving CQ obtained by the meta-analysis of the studies by Zhou et al. (Lancet, 2020) and Chen et al. (BMJ, 2020) (167/990=16.9%; 95% CI 14.5- 19.2).

**Figure 3.**
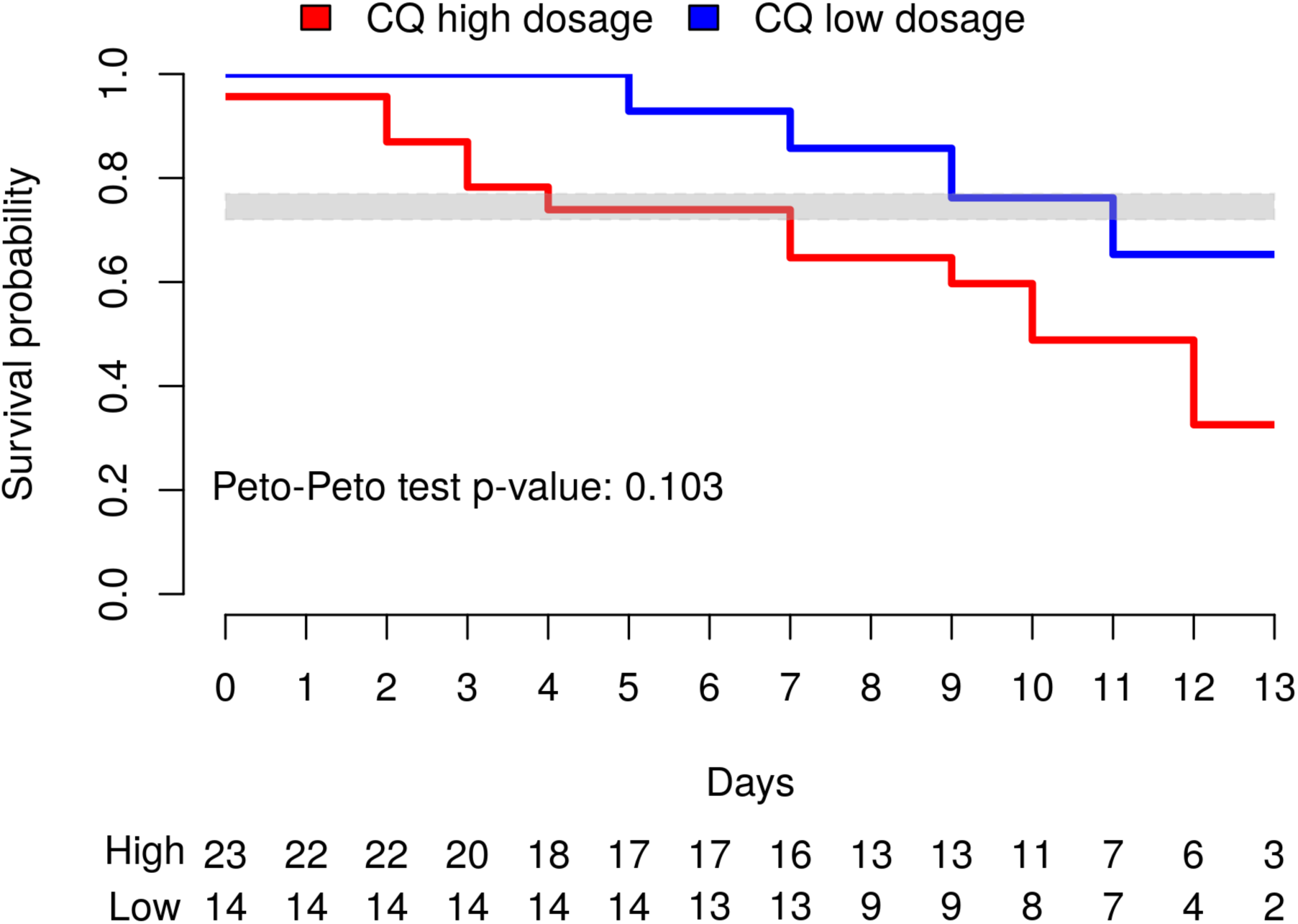
Time (in days) from randomization to death, in patients treated with each chloroquine dosage in a subgroup patients enrolled already in ICU. The gray band represents the upper and lower limits of the confidence interval for lethality in critically-ill patients not receiving CQ in the Italian cohort by Onder et al. (JAMA, 2020) (405/1,581=26%; 95% CI 23.5-27.8).

Based on the findings, DSMB recommended the immediate interruption of the high dosage arm and that all patients in it were unmasked and reverted to the low dosage arm.

Per protocol analysis was not performed due to the impossibility to monitor drug administration twice a day at the hospital. Radiological findings were presented in this manuscript only in the baseline due to the inability to perform careful analyses of the available CT scans over time. Radiological and complete efficacy data will be presented later.

## Discussion

In a unique pandemic situation, health professionals have to choose between offering medical assistance and generating and reporting reliable data, a dichotomy that compromises the generation of good quality evidence for clinical management. Global recommendations for COVID-19 are being made based on unpowered studies, however, and due to the chaotic urgency, such drugs are being prescribed in a compassionate manner given the severity of this disease. However, CQ, despite being a safe drug used for more than 70 years for malaria, might be toxic in the dosages recommended by Chinese authorities (high dosage 10g, for 10 days). Our study raises enough red flags to stop the use of such dosage (12g of CQ in total, for 10 days, due to the presentation of CQ tablets, 150mg, from *Farmanguinhos*) worldwide in order to avoid more unnecessary deaths. We were not able to independently assess the toxic role of azithromycin because all patients were already using this antibiotic as per hospital protocol. Oseltamivir, which also increases QTc, could potentiate cardiac side effects, because most of the patients (89.6%) were also in use of this drug for suspected influenza infection.

With the ethical impossibility of using a placebo arm, we were compelled to use historical data, based on very similar patients not using CQ. Fatality rates observed here were not lower, however one cannot reliably conclude that CQ is of no benefit. Placebo-controlled studies are being performed in countries not routinely using the drug. Several ongoing trials (including *CloroCovid-19 II* trial, from our group - <u>ClinicalTrials.gov</u>, number NCT04342650) have also been addressing the early use of CQ, in which the anti-inflammatory properties could potentially be more helpful. That information is urgently needed in placebo-controlled double-blinded randomized trials.

In addition to helping patients improve, CQ could be used to decrease the viral load in respiratory secretions, allowing less nosocomial and post-discharge transmission. However, our data provided no evidence of such an effect. Patients using CQ (irrespective of dosage) failed to present evidence of substantial viral clearance by the fifth day (day 4) of positive RT-PCR, even with the concomitant use of azithromycin.

CQ is recommended for the treatment of malaria in particular due to its low cost; few doses resulting in safe concentrations are needed to treat the disease^25^. CQ can deposit in tissues, especially the eye, causing retinal toxicity, which is associated only with prolonged use^23,24^. QTc prolongation >500ms was seen in 15.1% of patients, which is similar from what has been reported in patients with COVID-19 using HCQ (11.0%)^36^. No difference was seen between the arms. Therefore, we suggest that lethality attributed to higher dosage could have causes other than cardiotoxicity. Myopathy has also been associated with CQ use^25^. In our study, one patient developed rhabdomyolysis, which was attributed to CQ, and the drug was withdrawn.

In two patients, myocarditis was suspected based on the CKMB elevation since the first day of hospitalization, suggesting myocarditis related to SARS-CoV-2 itself. In such cases, drugs prolonging QTc could lead to severe arrhythmias. Unfortunately, this study’s randomization, probably due to the low sample size, assigned older patients with heart disease to the high dosage arm. Therefore, one limitation for the conclusions of the study on lethality per arm is that high CQ dosage arm presented more patients prone to cardiac complications, with or without CQ. In any case, the use of CQ in elderlies with heart disease should be made with caution.

Lethality in critically-ill patients seemed to be even higher than similar patients in Italy. That could reflect the quality of ICU in both countries or the possible lack of or deleterious effect of CQ in such patients with COVID-19.

The occurrence of myocarditis in our sample together with the confirmed QTc prolongation, warrants caution in relation to this drug’s safety, particularly considering the eventual increase in fatal arrythmias, such as ventricular tachycardia.

This study had some strengths, as it was: (1) double-blinded; (2) performed in a public hospital, which will represent most of the cases in countries like Brazil; (3) compliant with good clinical practices, with a vigilant and highly involved DSMB; (4) an assessment of two dosing schemes of CQ for the first time in COVID-19 patients.

Major limitations however included: (1) one single center; (2) not using a placebo control group as the use of placebo in Brazil in severe cases of COVID-19 infections is not considered ethically acceptable by national regulatory health agencies, especially due to the compassionate use of CQ – and because early reports seem to indicate its effectiveness *in vitro* and *in vivo*.

In conclusion, high CQ dosage scheme (12g), given for 10 days, was not sufficiently safe to warrant continuation of that particular study arm. We therefore strongly recommend that this dosage is no longer used anywhere for the treatment of severe COVID-19, especially because in the real world older patients using cardiotoxic drugs should be the rule. No apparent benefit of CQ was seen regarding lethality in our patients so far, but we will still enroll patients in the low CQ dose group to complete the originally planned sample size.

In order to better understand the role of CQ or HCQ in COVID-19, we recommend the following next steps: (1) trials evaluating its role as a prophylactic drug; (2) trials evaluating its efficacy against progression to severity when administered to patients with mild/moderate disease. Even if we fail to generate good evidence in time to control the current pandemic, the information will highly impact the way we deal with next coronavirus outbreaks in the future.

## Data Availability

All datas are available

## Contributors

FFAV, GCM, LAH, WMM, MVFG and MVGL conceived the study and developed the protocol with input from MGSB, VSS, RCP, DCBS, MPGM and QB. MGSB, MAAA, AMS, MPGM and MVGL supervised clinical work. GCM and FGN supervised laboratory work. MGSB supervised pharmacy work. VSS, LAH, DCBC, MSX, AS, JHRC, MLN, GASR, CJF, BCA, CTDR and AASB performed data management and analysis. VSS, JDBS, DCBS, AMS, QB, WMM, and MVGL wrote the manuscript with input from all other authors. CloroCovid-19 Team collected all the data. All authors critically read the manuscript and approved the final submitted version.

## Acknowledgments

Special thanks to the Brazilian Minister of Health Luiz Henrique Mandetta, to his Secretary of Health Surveillance (SVS) Wanderson Kleber Oliveira, and his Secretary of Science and Technology (SCTIE) Denizar Vianna, for all the remote support, and to Camile Sachetti (DECIT) and Jorge Souza Mendonça, director of Farmanguinhos. AMS and CTDR are fellows from *Fundação de Apoio à Pesquisa do Estado do Rio de Janeiro* (FAPERJ). FGN, JHRC, MLN, CTDR, WMM and MVGL are research fellows from *Conselho Nacional de Desenvolvimento Científico e Tecnológico* (CNPq). Special thanks to the director of *Hospital e Pronto Socorro Delphina Rinaldi Abdel Aziz*, José Luiz Gasparini, to the Governor of the Amazonas State, Wilson Lima, and to his Secretary of Health, Rodrigo Tobias, and Brazilian Senators Eduardo Braga and Eduardo Gomes. To the dean of *Universidade do Estado do Amazonas*, Cleinaldo Costa, for providing high-quality locally-produced personal protective equipment to the team. To the many industries from the Industrial Pole in Manaus (*Zona Franca de Manaus*), which donated tablet computers to the team for data collection at bedside, including business executives Wilson Périco and Marcelo Dutra. Many members from the CloroCovid-19 Team are funded by *Fundação de Amparo à Pesquisa do Estado do Amazonas* (FAPEAM) and CAPES. To ISGlobal, which receives support from the Spanish Ministry of Science and Innovation through *Centro de Excelencia Severo Ochoa 2019-2023* Program (CEX2018-000806-S), and support from *Generalitat de Catalunya* through the CERCA Program. CISM is supported by the Government of Mozambique and the Spanish Agency for International Development (AECID). We also thank Judith Recht and Donald Skillman for their thoughtful input on manuscript preparation.

